# Natural Language Processing for Automated Annotation of Medication Mentions in Primary Care Visit Conversations

**DOI:** 10.1101/2021.03.29.21254488

**Authors:** Craig H Ganoe, Weiyi Wu, Paul J Barr, William Haslett, Michelle D Dannenberg, Kyra L Bonasia, James C Finora, Jesse A Schoonmaker, Wambui M Onsando, James Ryan, Glyn Elwyn, Martha L Bruce, Amar K Das, Saeed Hassanpour

**Author notes:** Corresponding Author: Saeed Hassanpour, PhD, Postal address: One Medical Center Drive, HB 7261, Lebanon, NH 03756, USA.

## Abstract

**Objectives:** The objective of this study is to build and evaluate a natural language processing approach to identify medication mentions in primary care visit conversations between patients and physicians.

**Materials and Methods:** Eight clinicians contributed to a dataset of 85 clinic visit transcripts, and ten transcripts were randomly selected from this dataset as a development set. Our approach utilizes Apache cTAKES and Unified Medical Language System (UMLS) controlled vocabulary to generate a list of medication candidates in the transcribed text, and then performs multiple customized filters to exclude common false positives from this list while including some additional common mentions of the supplements and immunizations.

**Results:** Sixty-five transcripts with 1,121 medication mentions were randomly selected as an evaluation set. Our proposed method achieved an F-score of 85.0% for identifying the medication mentions in the test set, significantly outperforming existing medication information extraction systems for medical records with F-scores ranging from 42.9% to 68.9%.

**Discussion:** Our medication information extraction approach for primary care visit conversations showed promising results, extracting about 27% more medication mentions from our evaluation set while eliminating many false positives in comparison to existing baseline systems. We made our approach publicly available on the web as an open-source software.

**Conclusion:** Integration of our annotation system with clinical recording applications has the potential to improve patients’ understanding and recall of key information from their clinic visits, and, in turn, to positively impact health outcomes.

## BACKGROUND AND SIGNIFICANCE

Forty to eighty percent of healthcare information is forgotten *immediately* by patients post visit.[1–4] Poor recall and understanding of medical concepts have been identified as significant barriers to self-management, a central component of the Chronic Care Model, resulting in poorer health outcomes. [5–7] These barriers are amplified in older adults with multimorbidity, [8–11] where reduced cognitive capacity, [12–14] low health literacy,[15,16] and complex treatment plans are common.[17–19] Older adults with multimorbidity account for 96% of Medicare expenditures, and in the absence of optimal self-management, they experience lower quality of life and greater functional decline.[10,11,20–26]

An after-visit summary, shared via a patient portal, is a common strategy to improve recall of visit information.[27– 29] However, summaries impose a significant burden on clinicians who must document the entire visit in terms that are understandable to patients, with low health literacy being common.[30,31] Alternatively, audio recordings can provide a full account of the clinic visit and are an effective modality—71% of patients listen to recordings and 68% share their recording with a caregiver.[32] Clinic recordings improve patient understanding and recall of visit information, reduce anxiety, increase satisfaction, and improve treatment adherence.[32–38] As patient demand for recordings increases,[39,40] a growing number of clinics across the U.S. are offering audio recordings of clinic visits, and a recent survey reveals that almost a third of clinicians in the U.S. have shared a recording of a clinic visit with patients. [41]

Yet, unstructured clinic recordings may overwhelm patients.[39,42] Advances in data science methods, such as natural language processing (NLP), can be used to identify patterns in unstructured data and extract clinically meaningful information. These methods have been used to predict hospital readmissions,[43] future radiology utilizations,[44] and characterization of significance, change, and urgency of clinical findings in medical records.[45–49] As such, we plan to develop a recording system for patients that applies NLP methods to unstructured clinic visit recordings.[50]

In this paper, we describe an approach to extract mentions of medication names in transcripts of clinic visit audio recordings. Annotating mentions of medications discussed during a clinic visit recording can provide added value to the audio-recorded health information. We use NLP to highlight medication mentions in transcripts of clinic recordings. These annotations can be utilized to index the audio and aid visit recall by enabling key visit information to be easily accessed. In addition, the indexed medical concepts can be linked to credible and trustworthy online resources. These resources would provide additional information about medications to aid in patient understanding. Such an approach could potentially increase patient self-management, and, when shared with caregivers, could increase their confidence in delivering care.

At the time of this work, no prior work focused on extracting medication information from clinic visit conversations and their transcriptions. There has been some work toward the extraction of medication names and also prescription-related attributes such as dosage and frequency from medical text (primarily clinical notes). These systems have mainly focused on the extraction of medication information from written clinical notes. In 2009, the Third i2b2 Shared-Task on Challenges in Natural Language Processing for Clinical Data Workshop focused on medication information extraction. The challenge was to extract and label medication-related terms (medication name, dosage, frequency, etc.) from discharge summaries.[51] Teams were given 696 summaries for development, and then 547 summaries were used for evaluation. Twenty teams submitted entries to the challenge, with the top result for annotating medication names being an F-score of 90.3% on the evaluation dataset, utilizing a combination of a rule-based approach with two machine learning models (conditional random field and support vector machine). This top approach also achieved an F-score of 90.81% on an internal test set of 30 clinical records when evaluated by the system’s authors.[52]

Since the 2009 i2b2 challenge, additional work has been done to improve medication information extraction methods. Sohn et al. developed Medication Extraction and Normalization (MedXN) to extract medication information and map it to the most specific RxNorm concept possible.[53] This group reported an F-score of 97.5% for medication name on a test set of 26 clinical notes containing 397 medications. In 2014, MedEx, the system with the second-best results in the i2b2 challenge, was reimplemented using Unstructured Information Management Architecture (UIMA) to extract drug names and map them to both generalized and specific RxNorm concepts.[54] This system, named MedEx-UIMA, achieved an F-score of 97.5% for extracting and mapping to the most generalized concept and an F-score of 88.1% for mapping to the most specific concept, evaluating on a set of 125 discharge summaries from the original i2b2 challenge. The authors concluded that the new MedEx-UIMA implementation was consistent with and sometimes outperformed the original MedEx method. Most recently, PredMed was developed to extract medication names and related terms from office visit notes.[55] The comparison of PredMed for extracting medication names to earlier versions of MedEx and MedXN on a test set of 50 visit encounter notes showed F-scores of 80.0% for PredMed, 74.8% for MedEx, and 83.9% for MedXN. Since MedEx-UIMA and MedXN are available as open source systems, we used these systems as baselines for comparison in our study.

In another related work, Kim et al. developed a method for retrieval of biomedical terms in tele-health call notes.[56] Their team identified two types of noise in these records, explicit – including “spelling errors, unfinished sentences, omission of sentence marks, etc.” – and implicit – “non-patient information and a patient’s untrustworthy information” – and sought to remove that noise as part of their method. Utilizing a bootstrapping-based pattern learning process to detect variations related to the explicit noise, and dependency path-based filters to remove the implicit noise, their system achieved an F-score of 77.33% for detecting biomedical terms on evaluation data from 300 patients. This tool and its corresponding code base in not publicly available for comparison for this study. Furthermore, recently, there has been additional work on the analysis of medical conversation.[57–61] However, the corresponding models and methodologies for these studies are not publicly available for evaluation and comparison in this project.

## Materials and Methods

Our NLP pipeline was developed and validated to extract medication mentions in clinic visit transcripts. We define medication mentions as any place in the text that a term refers to a medication by a specific or general name or common lay term. Our pipeline takes advantage of Apache clinical Text Analysis and Knowledge Extraction System (cTAKES)[62] to generate a primary candidate list of medication mentions. Subsequently, our approach filters out false positive medical mentions in this list and adds the medication mentions that cTAKES misses in visit transcripts.

### Visit transcripts dataset

Transcripts of 85 patient visits with a primary care physician were used as our dataset in this study. These visits were audio recorded and transcribed by a HIPAA compliant commercial medical transcription service. These recordings, which came from eight clinicians, were 31 minutes long on average, ranging from 5.5 to 70.5 minutes. This study and the use of human subject data in this project were approved by the committee for the Protection of Human Subjects at Dartmouth College (CPHS STUDY#30126) with informed consent. Table 1 shows the demographics of the participants who had their clinical visit recordings used in our study.

**Table 1.** Participant demographics for transcribed visit recordings (SD: standard deviation).

|  | **Development Dataset** | **Validation Dataset** | **Evaluation Dataset** | **Total** |
| --- | --- | --- | --- | --- |
| Number of Recordings in Dataset | 10 | 10 | 65 | 85 |
| Participants with Demographic Data* | 9* | 10 | 54* | 73* |
| Gender |  |  |  |  |
| Female | 4 | 6 | 34 | 44 |
| Male | 5 | 4 | 20 | 29 |
| Mean age (SD) [Range] | 50.00 (18.57)<br>[23-87] | 58.60 (18.95)<br>[20-77] | 54.65 (15.61)<br>[25-92] | 54.62 (16.35)<br>[20-92] |
| Race |  |  |  |  |
| White | 9 | 10 | 54 | 73 |
| Ethnicity |  |  |  |  |
| Not Hispanic or Latino | 9 | 10 | 52 | 71 |
| Declines to list | - | - | 2 | 2 |
| Language Spoken |  |  |  |  |
| English | 9 | 10 | 54 | 73 |
| Recording length (SD) [Range] | 36.46 (17.37)<br>[17.55-70.39] | 37.07 (10.00)<br>[20.16-49.41] | 28.36 (11.95)<br>[5.42-55.33] | 30.55 (12.85)<br>[5.42-70.39] |
| Visit type |  |  |  |  |
| Annual physical established patient | 3 | 6 | 8 | 17 |
| Established patient follow-up | 2 | 3 | 29 | 34 |
| Same day add-on | 2 | 1 | 11 | 14 |
| New patient workup | 2 | - | 1 | 3 |
| History and physical | - | - | 2 | 2 |
| Other# | - | - | 3 | 3 |

\*Demographic data was not captured for 12 of the 85 transcripts.

Ten transcripts were randomly selected from this dataset as a development set. Another ten of the visit transcripts were randomly selected as a validation set for our model. The remaining 65 transcripts were reserved as a held-out test set for evaluation.

### Annotation for medication mentions

All the transcripts were independently annotated for medication mentions by two second-year medical students using the Extensible Human Oracle Suite of Tools (eHOST) software. [63] The two annotators initially worked through blocks of 5 or 10 transcripts, meeting after annotating each block to track inter-annotator agreement (IAA) on the identified medication mentions, discuss disagreements, and improve their accuracy in this annotation task, which led to steadily higher IAA over time. Our IAA calculation considers overlapping annotations as a match, allowing a flexible annotation arrangement for compound medication names. Once the annotators reached over 80% IAA, we considered them trained in this annotation task. Subsequently, they annotated the entire set of transcripts. Inter-annotator agreement for medication mentions between our annotators for the 65 transcripts in the evaluation dataset was 84.6%. In that dataset, Annotator 1 annotated 1,076 instances of medication mentions, and Annotator 2 annotated 1,048 instances of medication mentions.

### cTAKES baseline for annotating medications in transcripts

Our baseline approach was to utilize Apache cTAKES[62] to identify the medication mentions in the transcripts. cTAKES is an open-source widely-used NLP system for biomedical text processing. As one of its NLP capabilities, cTAKES is able to annotate and extract medical information from the free text of clinical reports. In its default configuration (i.e., Default Clinical Pipeline), cTAKES identifies parts of speech (POS) in input sentences, and annotates concepts found in the noun phrases that match with concepts from the Unified Medical Language System (UMLS) Metathesaurus.[64] We utilized the Default Clinical Pipeline of cTAKES (version 4.0.0) and its UMLS fast dictionary lookup functionality. The only modification to the default cTAKES configuration was to utilize its PrecisionTermConsumer function, which refines annotations to the most specific variation (e.g., if it finds the text “colon cancer” in a report, it only annotates “colon cancer” but not “colon” nor “cancer”). Since cTAKES is designed to work with medical record free text, there is an assumption that input text is a clinical note, written by an individual with a medical background. In contrast, the visit transcripts are typically a dyadic conversation between a patient and their physician.

### Our model for annotating medications in transcripts

After initial experiments with cTAKES and UMLS as a means to find medications mentioned in transcribed clinic visit conversations, we explored additional methods to filter out common false positives from the output generated by cTAKES. For this purpose, we took an iterative approach, looking at the most common errors in cTAKES outcomes for identification of medication mentions in our development set and developed new rule-based filters to detect and remove those from the cTAKES output. As our accuracy on the development set improved by filtering out many types of false positives (described in detail below), we ran our model against our validation set, finding that immunizations along with herbs and supplements persisted as typical errors. cTAKES had difficulty differentiating immunizations from diagnoses (e.g., chickenpox vaccine vs. chickenpox). Also, cTAKES did not annotate some commonly used herbs and supplements. In the next sections, we describe how our approach adds annotations for immunizations, herbs, and supplements, while filtering out false positives for medication mentions.

An overview of this approach is shown in Figure 1. We have made our code for this approach publicly available on GitHub.^1^

**Figure 1.**
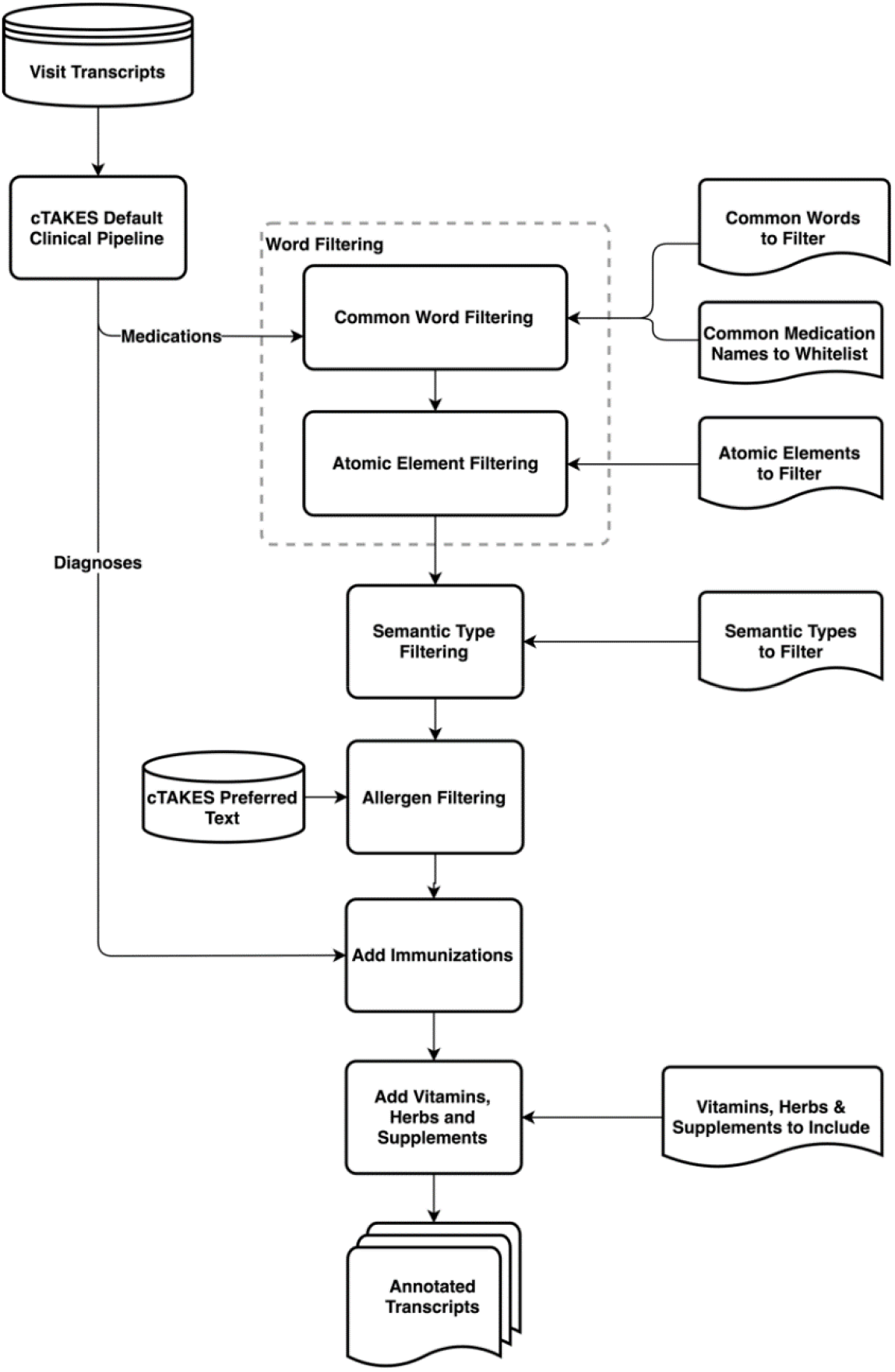
Overview of our approach to annotate medication mentions in clinic visit transcripts.

#### Common Word Filtering

Since many of the words appearing as false positives in the cTAKES output for medication annotations are common conversational words that have second meanings as medication names or acronyms (for example, “today” is also ToDAY, a name for an antibiotic primarily in veterinary use that appears in UMLS), we decided to utilize a large dictionary of common words to filter out these occurrences. We chose to use a dictionary of the 10,000 most common English words from Google’s Trillion Word Corpus^2^.[65] If any of those 10,000 words were annotated by cTAKES as a medication, our model removes that annotation, with a small subset of exceptions. From the 10,000 common words list, there were 24 words that are considered as exceptions and are allowed to remain annotated as medications. These words fit into three categories: 1) names of common medications (e.g., “Ambien”, “Insulin”, etc., which accounted for 17 of the 24); 2) generic terms (e.g., “herb”, “supplement”, and “vitamin”, along with their plurals); and 3) the word “flu”, which can refer to either a diagnosis or an immunization.

#### Chemical Element Filtering

cTAKES also annotates all chemical elements as medication mentions; “gold”, for example, can also be taken as a medication. In our approach, we systematically remove annotations for those chemical elements that are not typically taken as a medication or as a supplement.

#### UMLS Semantic Type Filtering

In our error analysis for cTAKES outputs, we also examined UMLS semantic types for the terms that cTAKES annotated as medication mentions. The six types shown in Table 2 generally produced false positives and few to no true positives. Our approach removes these sematic types as medication annotations from the cTAKES output where they occur.

**Table 2.**
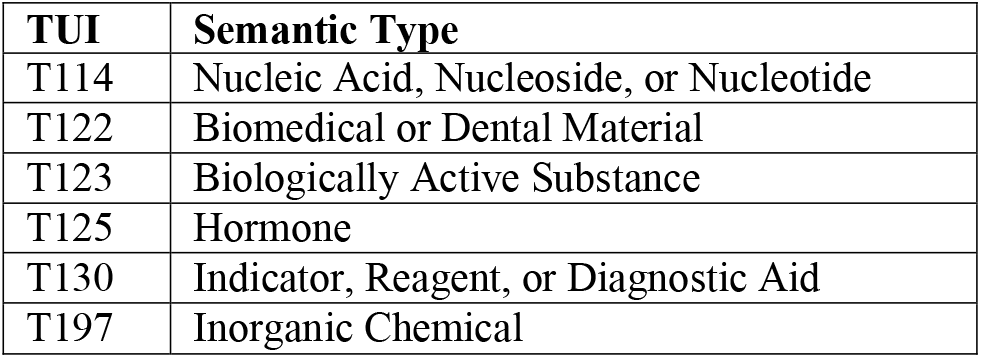
UMLS semantic types in cTAKES annotations that are filtered out in our approach.

#### Allergen Filtering

cTAKES annotates a number of food and food ingredient related terms (for example, “coconut”) as medication mentions, denoting them as an allergenic. We identify those annotations that have the word “allergenic” included in their preferred cTAKES text metadata, and we remove those annotations from the output of cTAKES in our model’s output.

#### Immunization Additions

A small number of medication-related UMLS terms are considered as both diagnoses and immunizations/vaccinations (for example, “flu” and “pertussis”). As a result, cTAKES annotation outputs were inconsistent about annotating these terms as immunizations/vaccinations or diagnoses. To improve the annotation of immunizations as medications, we also investigated the cTAKES diagnosis annotations. Since cTAKES segments the input text into sentences, we searched for the words “vaccine”, “shot”, “booster” and “pill” in the same sentence as a diagnosis annotation, and if both co-occurred, we annotated the diagnosis text as a medication.

#### Vitamin, Herb, and Supplement Additions

cTAKES also produces inconsistent results for annotating herbs and supplements. Our approach adds an additional dictionary of common herbs and supplements from MedlinePlus^3^ to capture these.[66]

### Evaluation

We applied our model on the evaluation dataset containing 65 transcripts to annotate medication mentions, in addition to capturing the original medication mention annotation output from cTAKES 4.0.0’s default clinical pipeline. We also applied publicly available MedEx-UIMA 1.3.7 and MedXN 1.0.1 software on the evaluation dataset to compare our results with their medication name annotations as the baselines.

For evaluation, we created a set of gold standard medication mentions in our evaluation dataset based on the work of our expert annotators. All medication mentions in our evaluation set that were agreed upon by the two expert annotators were kept in this gold standard set. A physician, trained in the method used by the annotators, served as an adjudicator to resolve disagreements between our annotators. A disagreement in the annotations would occur when one annotator had annotated a medication mention while the other had not. Disagreements were resolved by the adjudicating physician either choosing to keep the annotation from a single annotator in the gold standard set, or choosing to reject it. The adjudicating physician also reviewed disagreements between the output from our model and the set of annotations from human experts. Again, the adjudicator could either choose to keep the annotation from either source in the gold standard set or choose to reject it. As a result, a small number of medication mentions that were missed by both annotators were thus added to our gold standard set. The resulting gold standard evaluation dataset contained 1,121 medication mentions.

## Results

We calculated the standard evaluation metrics of precision, recall, and F-score for our proposed approach and the baseline methods using the medication mention gold standards in our evaluation set. We compared the results from cTAKES, MedEx-UIMA, MedXN, and our proposed model for identification of the gold standard medication mentions for the 65 transcripts in the evaluation set. Table 3 shows this comparison.

**Table 3.** The comparison of our proposed approach to existing baseline models for identification of gold standard medication mentions in our evaluation set.

| Model | # True Positives | # False Positives | # False Negatives | Precision | Recall | F-Score |
| --- | --- | --- | --- | --- | --- | --- |
| cTAKES | 1,119 | 2,814 | 163 | 28.5% | **87.3%** | 42.9% |
| MedEx-UIMA | 830 | 1,215 | 292 | 40.6% | 74.0% | 52.4% |
| MedXN | 832 | 318 | 432 | 72.3% | 65.8% | 68.9% |
| Our Approach | 1,062 | 168 | 206 | **86.3%** | 83.8% | **85.0%** |

## DISCUSSION

Our results indicate that the proposed approach significantly reduced the number of false positives, with a relatively small drop in the number of true positives and false negatives, in comparison to the best of three baseline models. As highlighted in Table 3, our proposed model has the best overall performance in comparison to the other baseline methods, with all of its evaluation metrics falling in the range of 83% to 87%. Overarching the finer aspects of our work is the observation that extracting medical terms from conversational dialogue between patients and their primary care physician has distinct challenges in comparison to extracting terms from typical clinical, note-like reports. To the best of our knowledge, the proposed work in this paper is the first attempt to extract medical terminology from conversations between a patient and their physician. Prior work for finding medication mentions has focused on written clinical reports.^47-51^

Our error analysis suggests that baseline approaches, which rely on dictionaries, struggle with patient-clinician conversational text because of language like filler words (for example, “aha” and “hmm”) matching with abbreviations for medications, and the fact that common conversational words are often used as medication names. We also observed, among the filters that we applied to the original cTAKES outputs, that filtering out “hormone” semantic type had the most impact on the improvement of the results. A slight but consistent majority of false positives in our dataset were from the discussion of lab test results, which will be a focus of our future work to improve the current results.

One advantage of our approach is that each portion of our pipeline was designed to generalize addressing issues seen during development, so our approach was able to recognize terms outside the development/validation datasets. Other rule-based and dictionary-based systems have often relied on whitelisting/blacklisting terms from their development datasets, which limits how they generalize outside their development data. For example, our use of the 10,000 most common English words from Google’s Trillion Word Corpus allows us to recognize and filter many common words. Our solutions to chemical elements, UMLS sematic types, allergens, immunizations, and vitamins/herbs/supplements were also all designed to potentially recognize terms outside what appeared in our development dataset.

Our evaluation has some limitations. Foremost, our evaluation dataset is relatively small and is from a single medical institution. We plan to extend our evaluation dataset in future work to test the generalizability of the proposed approach. In addition, because our gold standard was created by reaching consensus between two medical annotators and carrying out our approach, it is possible that other baseline methods, such as cTAKES, found a small number of true positives that were not accounted for by any of the annotators or our proposed method. That said, the sheer number of false positives generated by cTAKES makes adjudication of its medication mention output impractical. Finally, our approach, which is based on controlled vocabulary and rule-based filtering, does not consider word context and the corresponding contextual semantics in different circumstances. Since one of our goals is using these annotations to index segments of clinic visit conversations for end-users to review post-visit, we plan to conduct future work with end-users to determine how these limitations may impact the usability of the system.

Of note, as we fine-tuned our model on the validation set, we observed that context words in a sentence can be critical in our task, for example, for determining mentions of immunizations/vaccinations. Our result suggests that although the dictionary- and rule-based methods can achieve a promising result (F-score = 85%) for identification of medication mentions in clinic visit conversations, additional improvements in this domain will be gained through considering contextual semantics and machine learning models, which our team will pursue in future work.

## CONCLUSION

In this work, we developed an NLP pipeline for finding medication mentions in primary care visit conversations. The proposed model achieved promising results (Precision = 86.3%, Recall = 83.8%, F-Score = 85.0%) for identification of medication mentions in 65 clinic visit transcripts in our evaluation set. Since this is a first-of-a-kind study with clinic visit transcripts, we compared our approach to three existing systems used for extracting medication mentions from clinical notes. This comparison shows our approach can extract about 27% more medication mentions while eliminating many false positives in comparison to existing baseline systems. Integration of this annotation system with clinical recording applications has the potential to improve patients’ understanding and recall of key information from their clinic visits, and, in turn, behavioral and health-related outcomes. We plan to explore this potential in future trials of our system.

## Data Availability

The dataset used in this study includes patient health identifiers and is not publicly available.

## ACKNOWLEGEMENTS

The authors would like to thank Sheryl Piper and Roger Arend for their input on the project and thank Lamar Moss for his feedback on the manuscript.

## Footnotes

1 https://github.com/BMIRDS/HealthTranscriptAnnotator

2 https://github.com/first20hours/google-10000-english

3 https://medlineplus.gov/druginfo/herb_All.html

